# Prognostic Value of the Neutrophil Percentage-to-Albumin Ratio for All-Cause Mortality in Critically Ill Patients with Acute Myocardial Infarction: A Retrospective Cohort Study Based on the MIMIC-IV Database

**DOI:** 10.1101/2025.10.13.25337933

**Authors:** Chunxu Song, Kaiwen Xiao, Nan Zhang, Jingtao Cao, Jiayi Guo, Jiayi Shi, Chao Liu

## Abstract

**Background:** Critically ill patients with acute myocardial infarction (AMI) face high mortality, and existing risk stratification tools are limited. The neutrophil percentage-to-albumin ratio (NPAR), reflecting both systemic inflammation and nutritional status, may serve as a simple prognostic marker.

**Methods:** Data were extracted from the MIMIC-IV (version 2.0) database. Adult AMI patients with a first ICU admission between 2008 and 2019 were included. NPAR was calculated within 24 hours of ICU admission and categorized into quartiles. Cox regression models assessed associations with all-cause mortality. Kaplan–Meier analysis evaluated survival differences. Predictive performance of logistic regression, random forest, and XGBoost models was compared by ROC curves and AUC, and model interpretability was assessed using SHapley Additive exPlanations (SHAP).

**Results:** A total of 928 patients were included. Higher NPAR was associated with older age, greater illness severity, and higher mortality at all time points (p < 0.001). In univariate and partially adjusted Cox models, NPAR was an independent predictor of mortality, though significance diminished in the fully adjusted model (HR = 1.01, 95% CI 1.00–1.03, p = 0.055). Kaplan–Meier curves showed significantly poorer survival in higher quartiles (log-rank p < 0.0001). Logistic regression yielded the best predictive performance (AUC = 0.740), outperforming random forest (0.722) and XGBoost (0.709). SHAP analysis revealed a nonlinear effect of NPAR, with the strongest impact at intermediate levels, and age modified its contribution.

**Conclusion:** Elevated NPAR is associated with increased mortality in critically ill AMI patients. As a readily available and low-cost biomarker, NPAR may aid early risk stratification, though further prospective validation is required.

## 1. Introduction

Acute myocardial infarction (AMI) remains one of the leading causes of cardiovascular disease mortality worldwide, with persistently high in-hospital and long-term mortality rates, particularly among critically ill patients ^[1,2]^. Despite significant advances in recent years in reperfusion strategies, antiplatelet agents, and statins ^[3–6]^, substantial disparities in prognosis persist among AMI patients, suggesting limitations in traditional risk stratification tools for critically ill populations ^[7–10]^. Therefore, identifying novel, simple, and reliable biological markers to enhance early risk assessment holds significant clinical importance ^[11]^.

Inflammatory responses and nutritional status both play pivotal roles in the development and prognosis of AMI ^[12–17]^. Neutrophils, as core effector cells in acute inflammation, exacerbate myocardial injury by releasing inflammatory mediators and reactive oxygen species, while interacting with platelets to promote thrombosis ^[9,18–20]^. Low albumin levels not only reflect malnutrition and hepatic impairment but also indicate chronic inflammation and diminished physiological reserve capacity ^[21–23]^. In recent years, some scholars have proposed the neutrophil percentage-to-albumin ratio (NPAR) as a composite indicator integrating inflammation and nutritional status, which has demonstrated certain prognostic value in heart failure, angina pectoris, and stroke ^[24–26]^. However, the predictive role of NPAR in critically ill patients with AMI remains to be systematically evaluated.

Based on this, the present study utilizes the large-scale public intensive care database MIMIC-IV to investigate the relationship between NPAR and all-cause mortality risk in critically ill AMI patients. Furthermore, machine learning methods are employed to evaluate its predictive performance and interpretability. We hypothesize that elevated NPAR levels are significantly associated with poor outcomes, and that NPAR, as a simple and readily available indicator, may provide new reference criteria for risk stratification and clinical management of critically ill AMI patients.

## 2 Methods

### 2.1 Data source and study population

This retrospective cohort study was conducted using data from the MIMIC-IV (Medical Information Mart for Intensive Care IV, version 2.0) database, which contains detailed clinical information of patients admitted to the intensive care units (ICUs) of the Beth Israel Deaconess Medical Center between 2008 and 2019. One author completed the required training and obtained access to the database. Since all data were de-identified, individual informed consent was waived ^[27,28]^.

A total of 73,181 ICU admissions recorded in MIMIC-IV were initially screened. Patients diagnosed with AMI were identified using ICD-9 and ICD-10 codes, yielding 8,112 eligible cases. Exclusion criteria were as follows: (1) multiple ICU admissions, only the first admission was retained (n = 1,235); (2) age < 18 years (n = 5); (3) ICU length of stay < 24 hours (n = 2,560); and (4) missing neutrophil percentage or serum albumin values within the first 24 hours of ICU admission (n = 3,384). After applying these criteria, 928 patients were included in the final analysis. **(Figure 1)**

**Figure 1.**
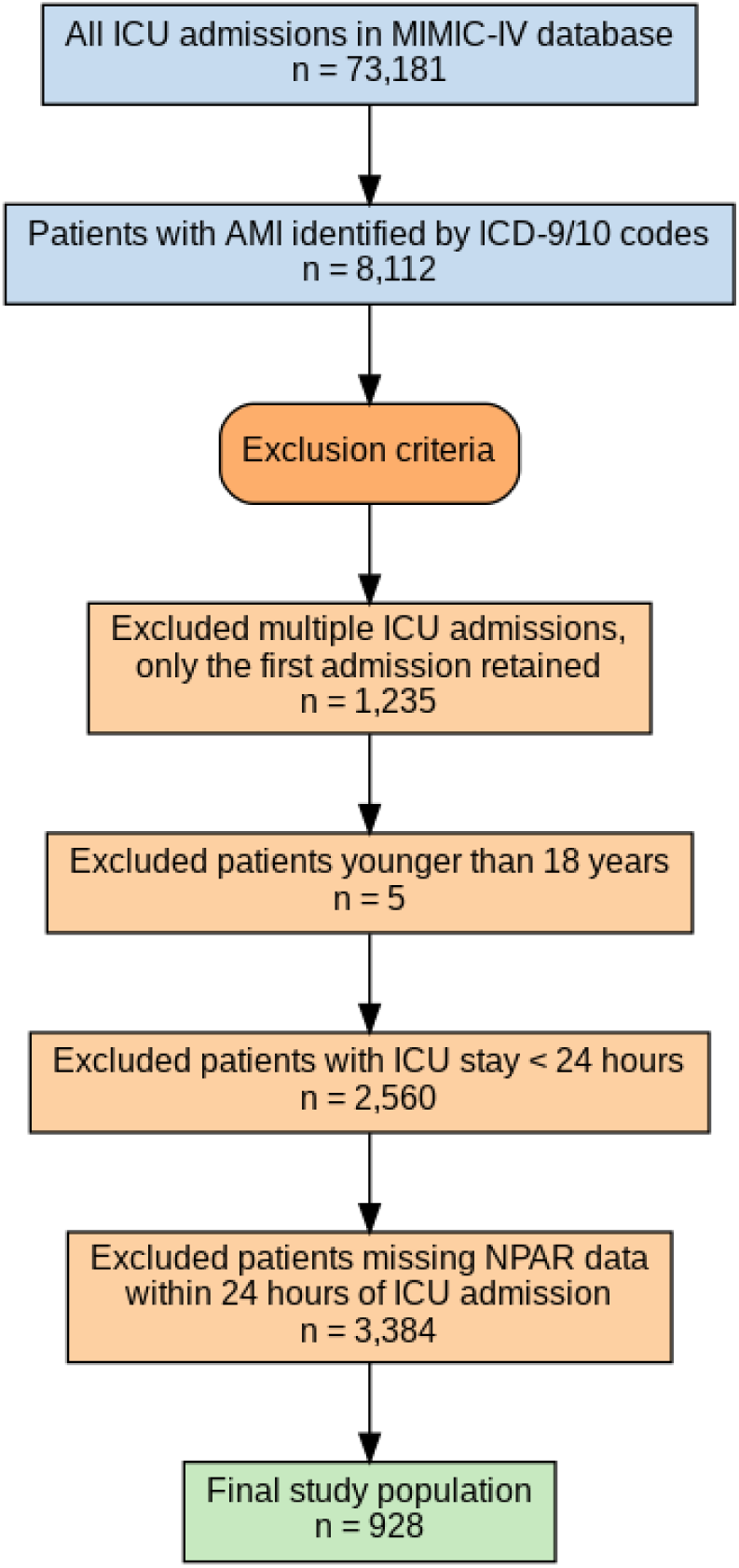
Flowchart of patient selection from the MIMIC-IV database.

### 2.2 Exposure variable and grouping

The exposure variable of interest was the NPAR, which was calculated as the ratio of neutrophil percentage (%) to serum albumin concentration (g/dL) measured within the first 24 hours after ICU admission. Based on the distribution of NPAR values, patients were categorized into four groups according to quartiles: Q1 (lowest), Q2, Q3, and Q4 (highest). This stratification was used to assess the association between increasing NPAR levels and clinical outcomes ^[29,30]^.

### 2.3 Outcomes

The primary outcome of this study was all-cause mortality, including in-hospital mortality, as well as 30-day, 90-day, 180-day, and 365-day mortality. Mortality status and dates of death were obtained from the in-hospital records and follow-up data contained in the MIMIC-IV database. Survival time was defined as the interval from the date of ICU admission to the date of death or the last follow-up. Patients who remained alive at the end of the observation period were censored ^[31,32]^.

### 2.4 Covariates

Baseline demographic variables included age, sex, and race. Laboratory parameters within the first 24 hours of ICU admission were extracted, including white blood cell count, hemoglobin, platelet count, red cell distribution width (RDW), prothrombin time (PT), serum creatinine, blood urea nitrogen (BUN), alanine aminotransferase (ALT), aspartate aminotransferase (AST), serum electrolytes (sodium, potassium), and blood glucose. Vital signs included heart rate, mean blood pressure (MBP), and respiratory rate. Disease severity was assessed using the Sequential Organ Failure Assessment (SOFA) score and Simplified Acute Physiology Score II (SAPS II). In addition, comorbidities such as coronary artery disease (CAD), congestive heart failure (CHF), atrial fibrillation (AF), chronic kidney disease (CKD), chronic obstructive pulmonary disease (COPD), hypertension, and diabetes were identified based on ICD-9/10 codes. Medication use during hospitalization, including aspirin, clopidogrel, β -blockers, angiotensin-converting enzyme inhibitors/angiotensin II receptor blockers (ACEI/ARB), and statins, was also recorded.

### 2.5 Statistical analysis

Continuous variables were expressed as mean ± standard deviation (SD) and compared among NPAR quartiles using one-way analysis of variance (ANOVA). Categorical variables were presented as counts and percentages, and differences were assessed using the chi-square test. Kaplan–Meier survival curves were plotted to compare survival probability across NPAR quartiles, with the log-rank test used to assess statistical significance. Cox proportional hazards regression models were constructed to evaluate the association between NPAR and all-cause mortality. Three models were built: Model 1 (univariate), Model 2 (adjusted for age and sex), and Model 3 (further adjusted for hemoglobin, creatinine, BUN, heart rate, SOFA score, SAPS II, CKD, COPD, CAD, and statin use). To further evaluate predictive performance, three machine learning models (logistic regression, random forest, and XGBoost) were constructed, and their discrimination ability was compared using receiver operating characteristic (ROC) curves and the area under the curve (AUC). Model interpretability was assessed using SHapley Additive exPlanations (SHAP) to quantify the contribution of NPAR and other variables to mortality risk prediction. All analyses were conducted using R software (version 4.2.2). A two-tailed p < 0.05 was considered statistically significant.

## 3 Results

### 3.1 Baseline Characteristics of Study Participants

A total of 928 critically ill patients with acute myocardial infarction were divided into NPAR quartiles. Compared with the lowest quartile, patients in higher NPAR groups were slightly older and showed significantly increased neutrophil percentage, white blood cell count, renal and liver function markers, as well as higher SOFA and SAPS II scores, while albumin and hemoglobin levels decreased (all p < 0.05). The prevalence of chronic kidney disease and COPD increased with higher NPAR, whereas coronary artery disease was less frequent. In addition, use of aspirin, β-blockers, ACEI/ARB, and statins declined across quartiles. Importantly, in-hospital, 30-day, 90-day, 180-day, and 365-day mortality all rose markedly with higher NPAR (all p < 0.001). **(Table 1)**

**Table 1.**
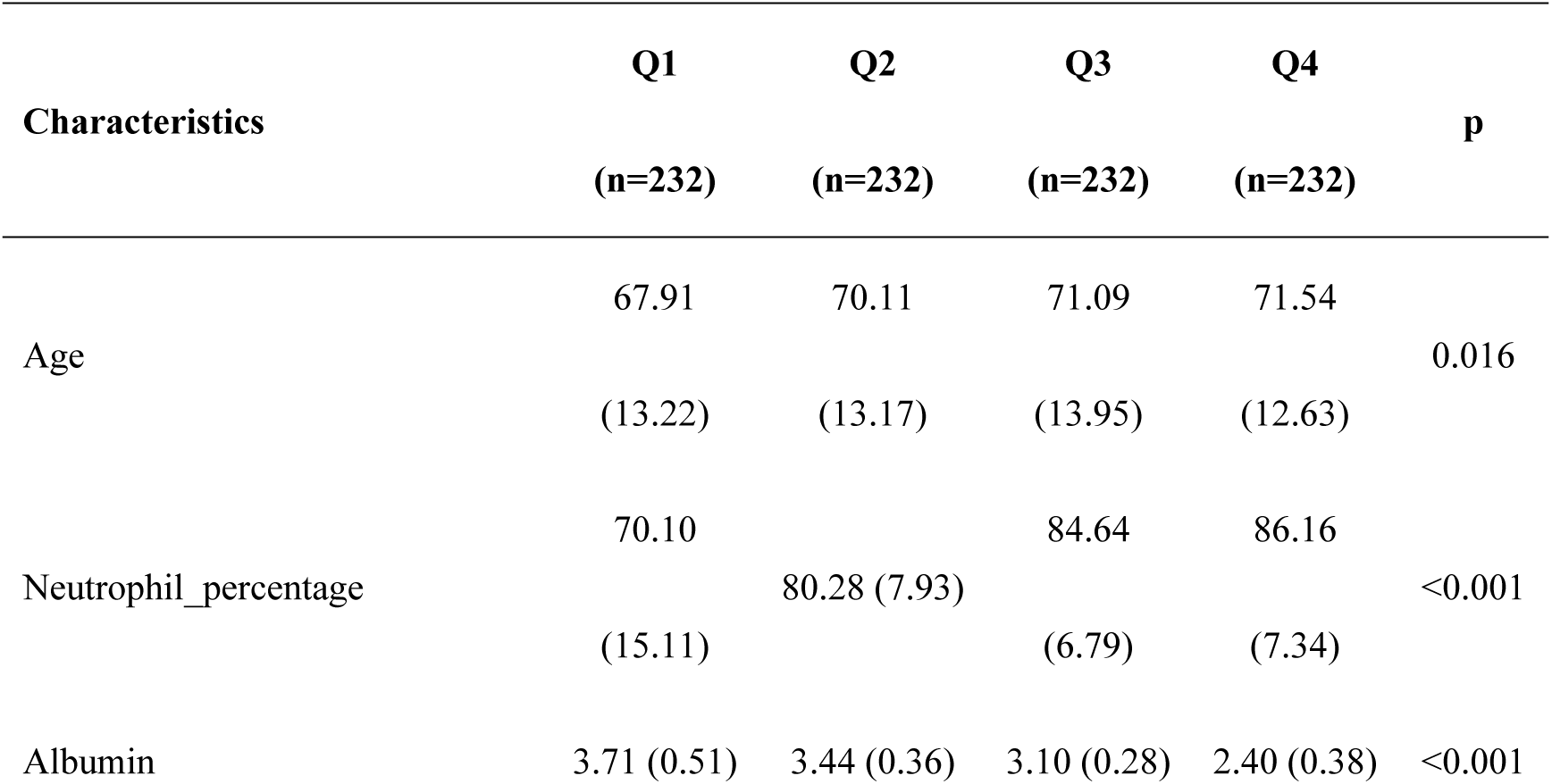
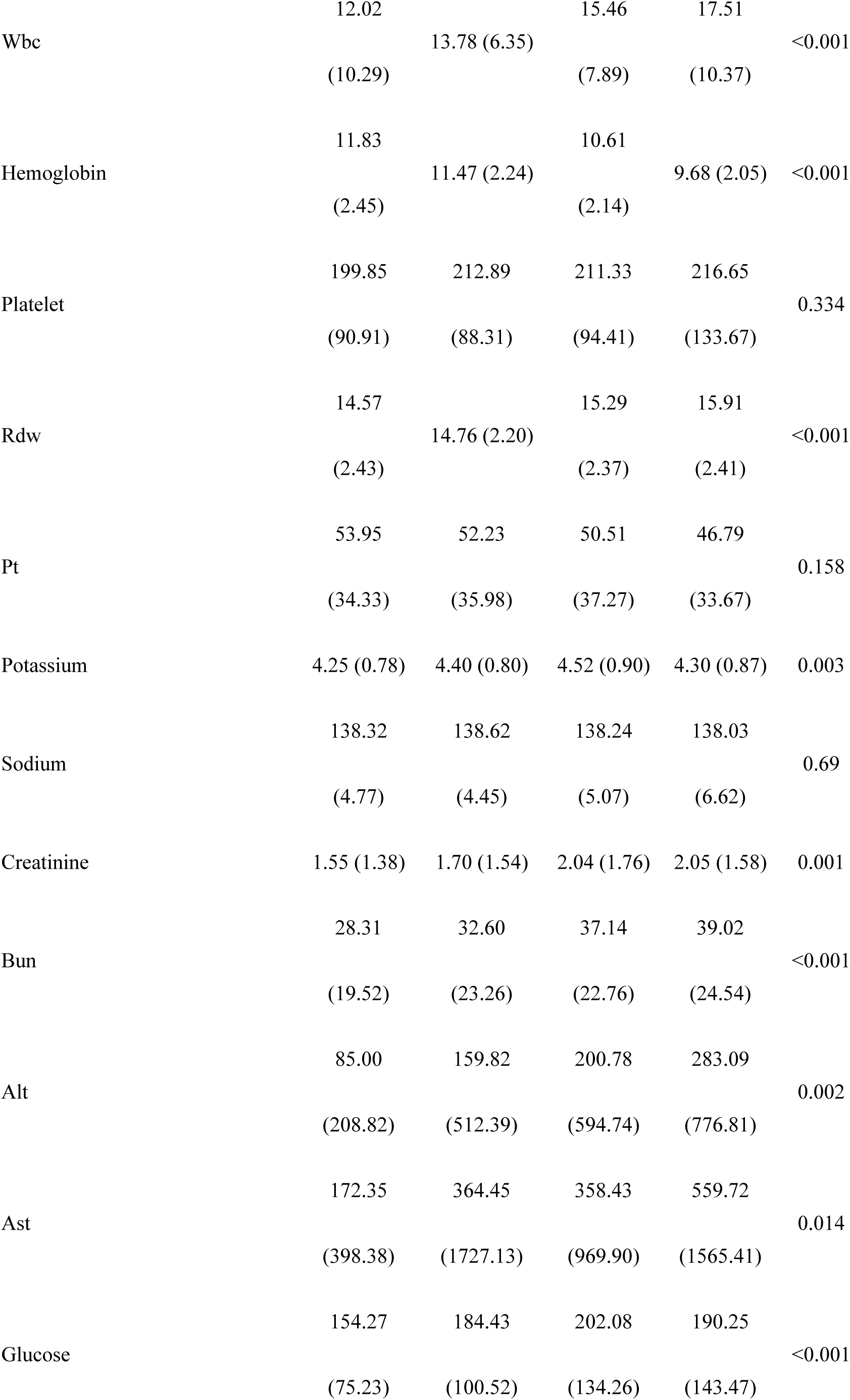
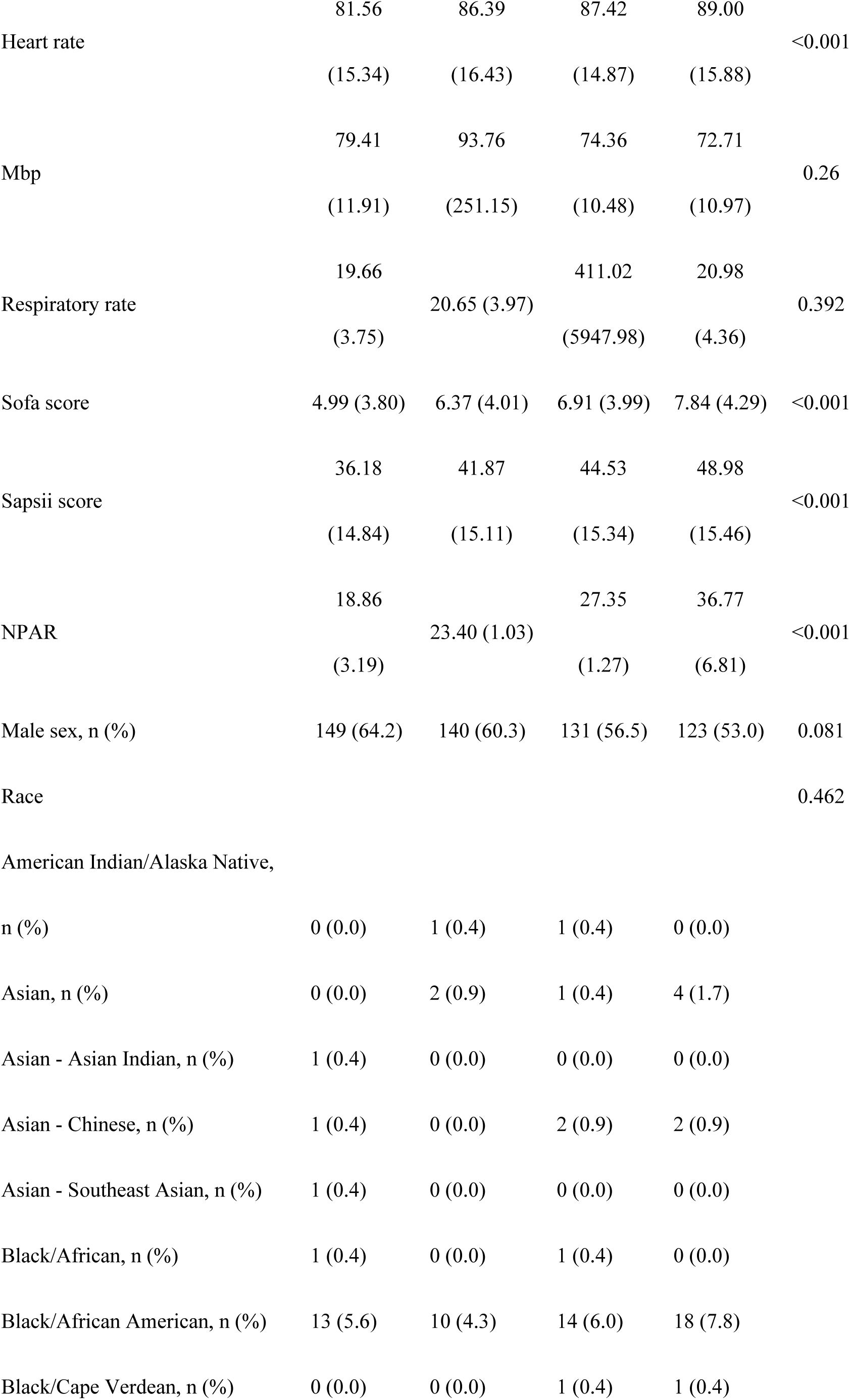
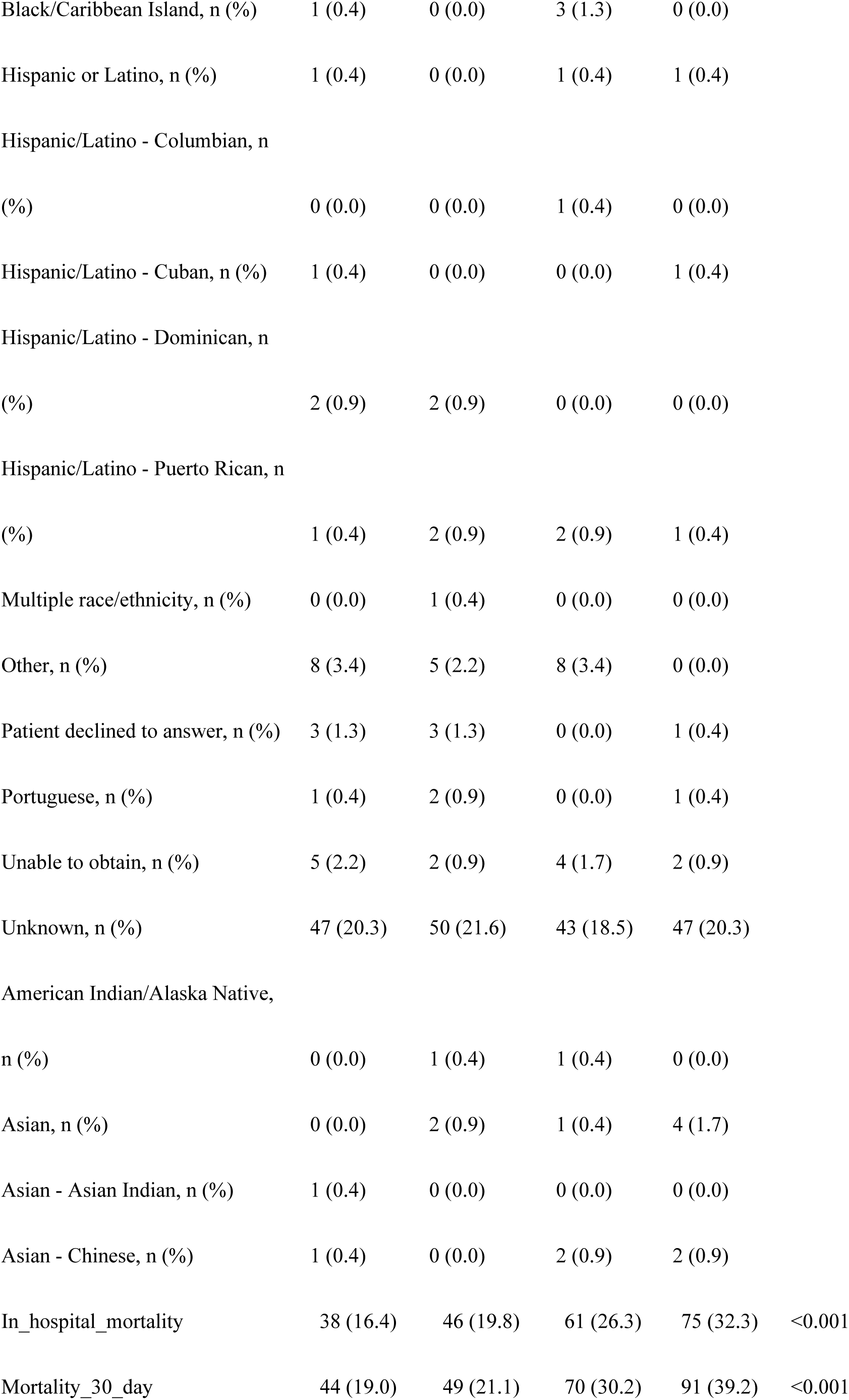
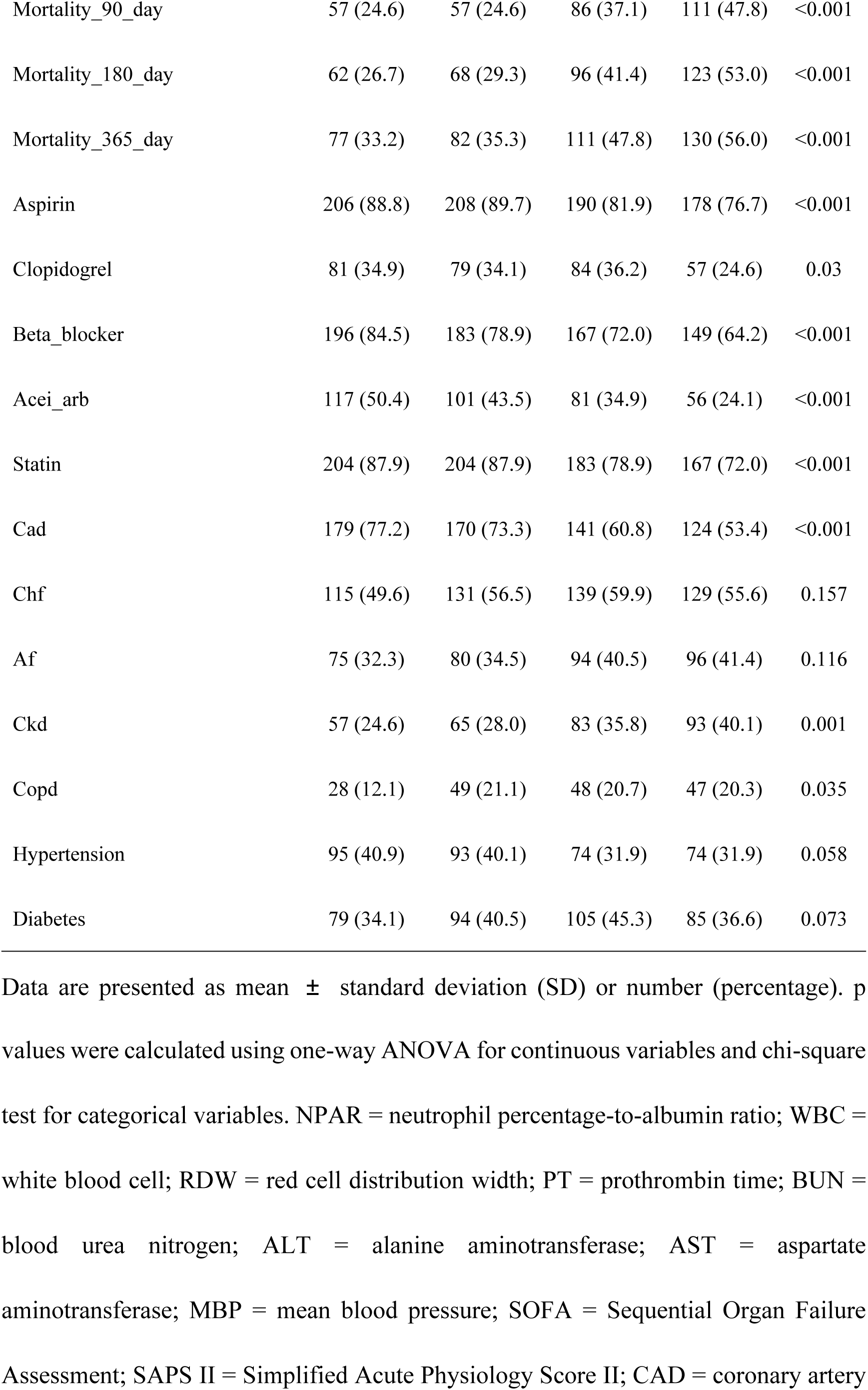
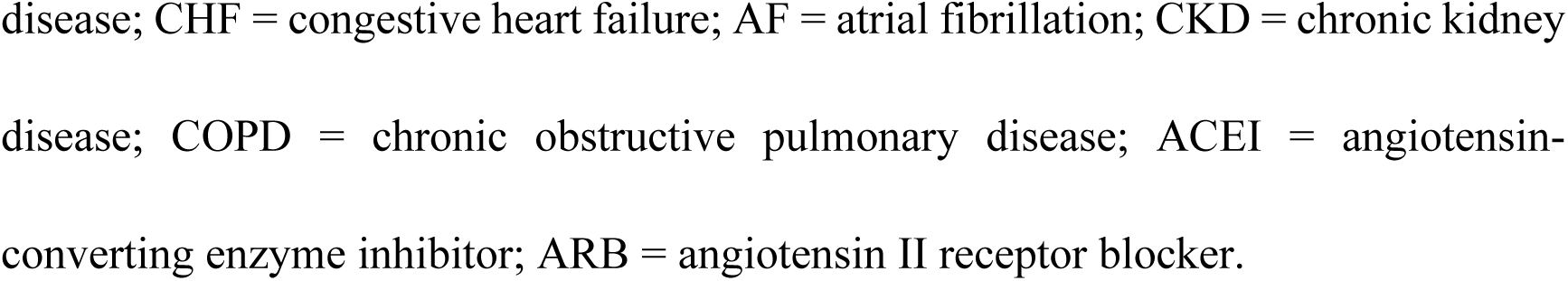
Baseline characteristics of critically ill patients with acute myocardial infarction stratified by quartiles of NPAR.

### 3.2 Association between NPAR and all-cause mortality

In the univariate analysis, higher NPAR was significantly associated with increased risk of all-cause mortality (HR=1.03, 95% CI 1.02–1.04, p < 0.001). This association remained robust after adjustment for age and gender (HR=1.03, 95% CI 1.02–1.05, p < 0.001). However, in the fully adjusted model, the association was attenuated and did not reach statistical significance (HR=1.01, 95% CI 1.00–1.03, p = 0.055). In the fully adjusted model, several covariates emerged as significant predictors of all-cause mortality. Lower hemoglobin (HR=0.91, 95% CI 0.87–0.96, p < 0.001), higher SOFA score (HR=1.06, 95% CI 1.02–1.11, p = 0.002), and higher SAPS II score (HR=1.01, 95% CI 1.00–1.02, p = 0.012) were independently associated with increased mortality risk. By contrast, creatinine, BUN, heart rate, CKD, COPD, CAD, and statin use were not significantly associated with mortality after adjustment. **(Table 2)**

**Table 2.** Cox proportional hazards regression analysis of NPAR and all-cause mortality in critically ill patients with acute myocardial infarction.

| Characteristic | Model 1 | p-value | Model 2 | p-value | Model 3 | p-value |
| --- | --- | --- | --- | --- | --- | --- |
|  | HR (95%<br>CI) |  | HR (95%<br>CI) |  | HR (95% CI) |  |
| NPAR | 1.03 (1.02- |  | 1.03 (1.02- |  |  |  |
| (continuous) | 1.04) | <0.001 | 1.05) | <0.001 | 1.01 (1.00-1.03) | 0.055 |
| Age at ICU | 1.03 (1.02- |  | 1.03 (1.02- |  |  |  |
| admission | 1.04) | <0.001 | 1.04) | <0.001 | 1.03 (1.02-1.04) | <0.001 |
| Gender: Female | Reference | — | Reference | — | Reference | — |
|  | 1.01 (0.83- |  | 1.01 (0.83- |  |  |  |
| Gender: Male | 1.23) | >0.9 | 1.23) | >0.9 | 0.95 (0.77-1.17) | 0.600 |
| Hemoglobin | — | — | — | — | 0.91 (0.87-0.96) | <0.001 |
| Creatinine | — | — | — | — | 0.99 (0.91-1.08) | 0.800 |
| BUN | — | — | — | — | 1.00 (1.00-1.01) | 0.500 |
| Heart rate | — | — | — | — | 1.00 (1.00-1.01) | 0.600 |
| SOFA score | — | — | — | — | 1.06 (1.02-1.11) | 0.002 |
| SAPS II score | — | — | — | — | 1.01 (1.00-1.02) | 0.012 |
| CKD | — | — | — | — | 1.13 (0.89-1.42) | 0.300 |
| COPD | — | — | — | — | 1.24 (0.98-1.56) | 0.079 |
| CAD | — | — | — | — | 1.04 (0.83-1.30) | 0.700 |
| Statin use | — | — | — | — | 0.86 (0.67-1.11) | 0.200 |

Model 1: univariate analysis.
Model 2: adjusted for age and gender.
Model 3: adjusted for age, gender, hemoglobin, creatinine, BUN, heart rate, SOFA score, SAPS II score, CKD, COPD, CAD, and statin use.
HR=hazard ratio; CI= confidence interval; NPAR = neutrophil percentage-to-albumin ratio; BUN=blood urea nitrogen; SOFA=Sequential Organ Failure Assessment; SAPS II=Simplified Acute Physiology Score II; CKD=chronic kidney disease; COPD=chronic obstructive pulmonary disease; CAD=coronary artery disease.

**Figure 2** illustrates the Kaplan–Meier survival curves for all-cause mortality across NPAR quartiles. Patients in the highest NPAR quartile (Q4) had the poorest survival, while those in the lowest quartile (Q1) demonstrated the most favorable prognosis. The survival probability decreased progressively with increasing NPAR levels, and the log-rank test indicated a highly significant difference among the four groups (p < 0.0001). **(Figure 2)**

**Figure 2.**
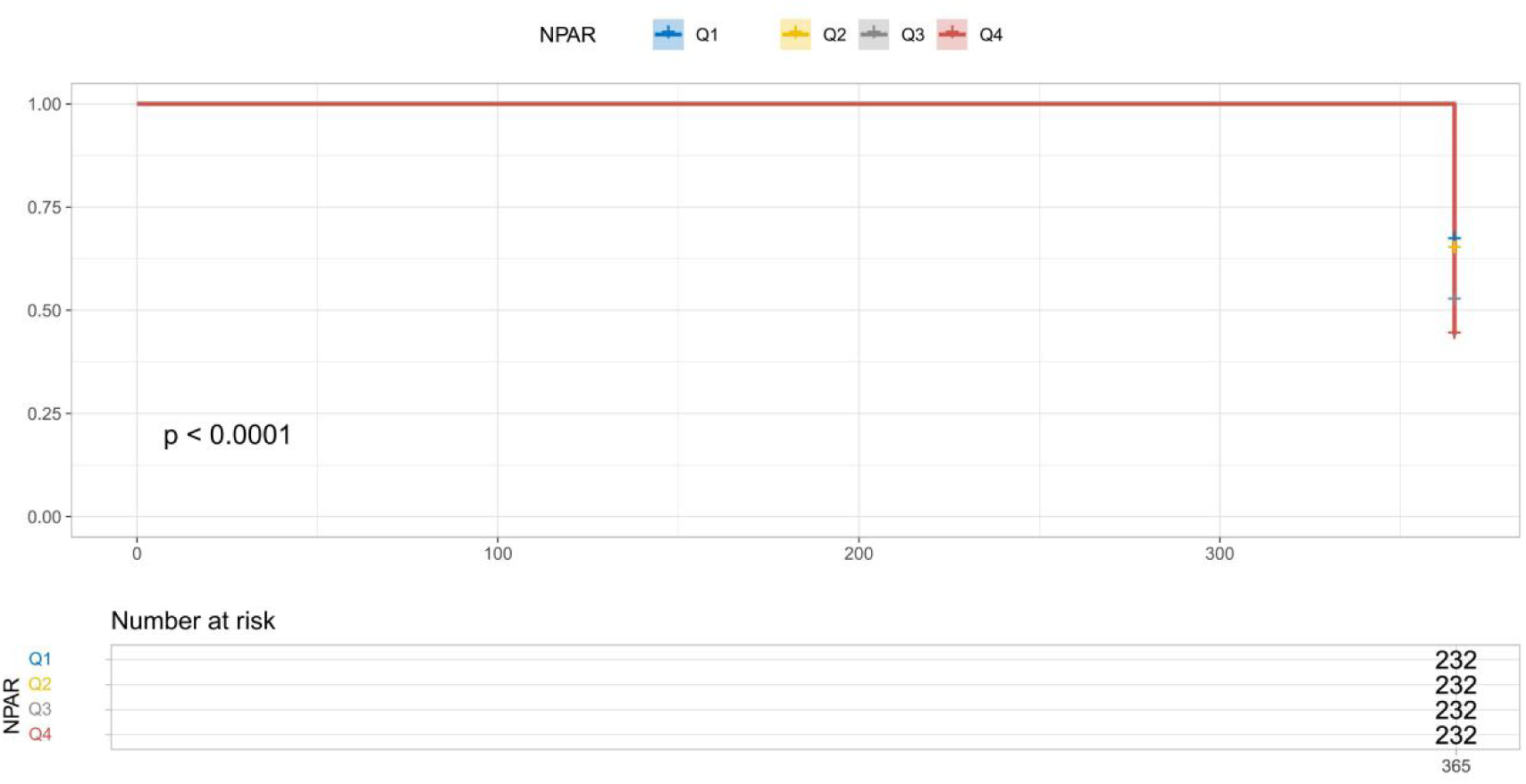
Kaplan–Meier survival curves for all-cause mortality according to quartiles of NPAR. Q1–Q4 represent increasing quartiles of NPAR, with Q1 as the lowest and Q4 as the highest.

### 3.3 Comparison of ROC curves among three machine learning models

The discriminative performance of the three machine learning models for predicting all-cause mortality in critically ill AMI patients was assessed by ROC curve analysis. Logistic regression achieved the highest AUC (0.740), followed by random forest (AUC = 0.722) and XGBoost (AUC = 0.709). Although all three models demonstrated acceptable predictive ability, logistic regression outperformed the other models in overall discrimination, while XGBoost showed the lowest AUC. (**Figure 3**)

**Figure 3.**
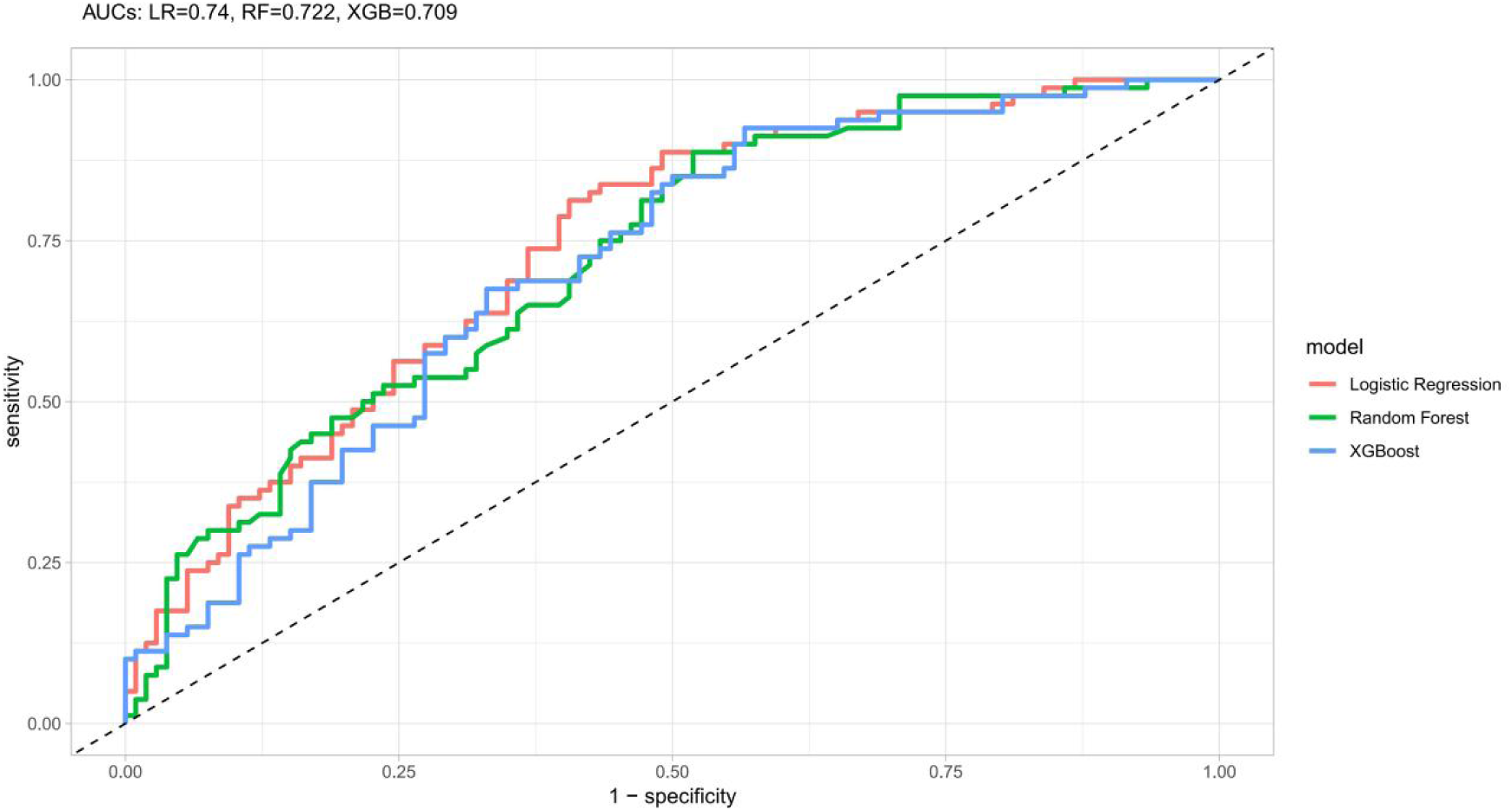
Comparison of receiver operating characteristic (ROC) curves of three machine learning models for predicting all-cause mortality in critically ill patients with acute myocardial infarction.

Figure 4 shows the SHAP dependence plot for NPAR. The curve demonstrated a nonlinear relationship between NPAR and its contribution to mortality risk prediction. NPAR exerted the strongest positive impact on risk at intermediate values, while both very low and very high NPAR levels were associated with relatively smaller contributions to predicted risk. In addition, the color distribution indicated that age may further modulate the effect of NPAR, with older patients showing higher SHAP values at comparable NPAR levels. **(Figure 4)**

**Figure 4.**
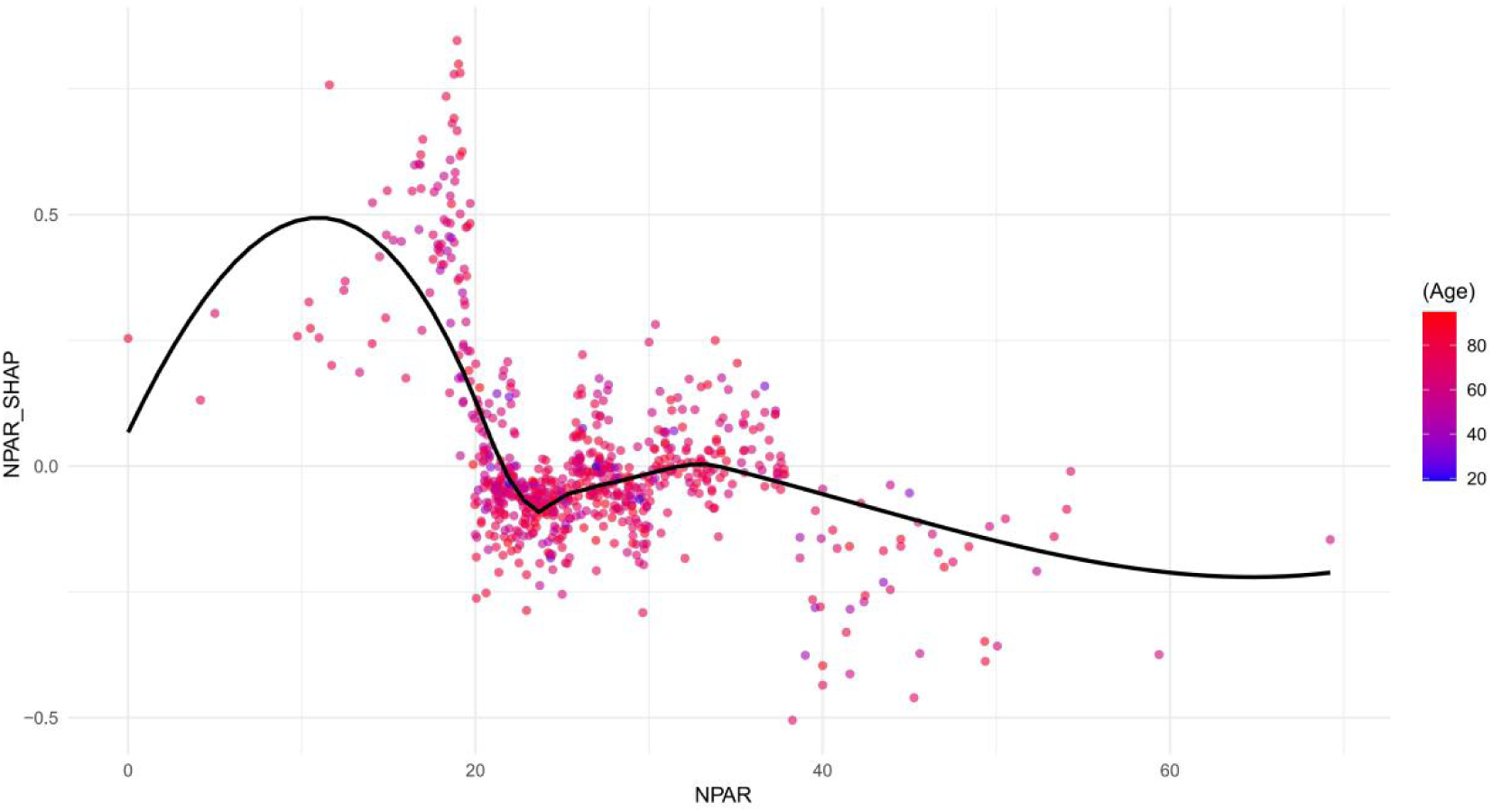
SHAP dependence plot for the association between NPAR and mortality risk in critically ill patients with acute myocardial infarction. Each point represents an individual patient, with color indicating age. Higher SHAP values correspond to stronger positive contributions to mortality risk.

## 4 Discussion

This study systematically evaluated the prognostic value of the NPAR in critically ill patients with AMI using the MIMIC-IV database. Results demonstrated that elevated NPAR was closely associated with worsening disease severity and significantly increased mortality. NPAR emerged as an independent risk factor in both univariate and partially adjusted models, though its effect diminished in the fully adjusted model, suggesting its prognostic role may be partially mediated by other clinical variables. Kaplan–Meier survival curves further confirm significantly reduced long-term survival rates in patients with high NPAR. Machine learning model comparisons reveal that logistic regression outperforms random forest and XGBoost in predicting all-cause mortality. SHAP dependency plots revealed a nonlinear relationship between NPAR and mortality risk, with moderate NPAR levels contributing most significantly to mortality prediction, while extremely low or high levels contributed relatively less. Age was identified as a modulating factor influencing this association.

This study found that NPAR was closely associated with indicators of disease severity (e.g., SOFA, SAPS II) and markers of inflammation and nutritional status (e.g., white blood cell count, hemoglobin, albumin) in critically ill AMI patients. As NPAR levels increased, both short-term and long-term mortality rates significantly rose. This aligns with previous findings that both inflammatory responses and hypoalbuminemia are strongly associated with poor outcomes in AMI patients ^[33–36]^. As a composite indicator integrating inflammation and nutritional status, NPAR may provide a more comprehensive reflection of patients’ systemic condition than individual markers, thereby demonstrating potential prognostic value.

Elevated NPAR levels may adversely affect AMI prognosis through the following mechanisms. First, neutrophils rapidly migrate to the ischemic myocardial region during the acute phase of AMI, directly exacerbating myocardial cell necrosis and apoptosis by releasing reactive oxygen species, proteases, and inflammatory mediators ^[37,38]^. Additionally, interactions between neutrophils and platelets promote thrombus formation, leading to expanded infarct size and exacerbated reperfusion injury ^[39]^. Studies have demonstrated that elevated neutrophil ratios are closely associated with infarct size, impaired cardiac function, and adverse cardiovascular events in AMI patients ^[40,41]^. Secondly, albumin not only serves as a sensitive indicator of nutritional status but also participates in multiple physiological functions, including anti-inflammatory and antioxidant activities, as well as maintaining capillary permeability ^[42,43]^. Hypoalbuminemia often indicates chronic inflammatory activation, impaired liver function, or malnutrition, potentially weakening the body’s ability to respond to acute myocardial injury. It is also associated with vascular endothelial dysfunction, immunosuppression, and coagulation abnormalities ^[44–48]^. This study found that the effect of NPAR weakened in the fully adjusted model, suggesting that its partial effect may be mediated by severity-of-illness scores (SOFA, SAPS II) and anemia levels. This indicates that NPAR is not an isolated influencing factor but interacts with multiple risk factors such as anemia, renal insufficiency, and systemic inflammatory response to jointly determine outcomes in AMI patients ^[49–51]^.

In the machine learning model comparison of this study, logistic regression achieved the highest AUC (0.740), outperforming both random forest (0.722) and XGBoost (0.709). This result suggests that traditional statistical models like logistic regression maintain robust stability and generalization capabilities even when dealing with limited data and strong variable correlations. Although ensemble algorithms demonstrate advantages in handling complex nonlinear relationships, they did not yield significant improvements in this cohort. This may be attributed to factors such as sample size, variable characteristics, and model tuning strategies.

Through the SHAP dependency plot, we further observed that NPAR exhibits a nonlinear pattern in influencing mortality risk, contributing most significantly at moderate levels while its effect diminishes at extremely low or high values. This may reflect distinct pathophysiological mechanisms across different clinical subgroups. Additionally, elderly patients demonstrate a more pronounced risk contribution under high NPAR states, suggesting that age serves as a critical modifier in how inflammation–malnutrition imbalance impacts mortality risk ^[52–54]^.

## 5 Limitations

There are several limitations in this study. First, the retrospective, single-center design introduces selection bias and limitations in causal inference. Second, NPAR was measured only upon ICU admission, lacking assessment of dynamic changes, which may underestimate its predictive value. Third, some variables had missing data or measurement errors; although statistically controlled, these may still influence the results. Fourth, the study population primarily consisted of American ICU patients, and the generalizability to other populations requires further validation. Fifth, the exploration of mechanisms was largely based on speculation, lacking direct evidence from experimental studies, and necessitates further basic and prospective clinical research for confirmation.

## 6 Conclusion

This study demonstrates that elevated NPAR levels are significantly associated with all-cause mortality risk in critically ill patients with acute myocardial infarction and possess certain risk stratification value. As a simple, low-cost indicator, NPAR holds promise for clinical prognosis assessment; however, its independent predictive role and underlying mechanisms require further validation through prospective studies.

## Data availability

The datasets analyzed in this study are publicly available in the MIMIC-IV database (version 2.0), which can be accessed at: https://physionet.org/content/mimiciv/2.0/. Access is available to qualified researchers who complete the necessary data use agreement and training.

## Acknowledgments

We thank the MIMIC-IV database for providing the original study data.

## Author Contributions

Data curation: Chunxu Song, Kaiwen Xiao, Nan Zhang, Jingtao Cao, Jiayi Guo, Jiayi Shi.

Writing – original draft: Chunxu Song, Kaiwen Xiao.

Writing – review & editing: Chao Liu

## List of Abbreviations

(AMI): acute myocardial infarction
(NPAR): neutrophil percentage-to-albumin ratio
(RDW): red cell distribution width
(PT): prothrombin time
(BUN): blood urea nitrogen
(ALT): alanine aminotransferase
(AST): aspartate aminotransferase
(MBP): mean blood pressure
(SOFA): Sequential Organ Failure Assessment
(SAPS II): Simplified Acute Physiology Score II
(CAD): coronary artery disease
(CHF): congestive heart failure
(AF): atrial fibrillation
(CKD): chronic kidney disease
(COPD): chronic obstructive pulmonary disease
(MIMIC-IV): Medical Information Mart for Intensive Care IV
(ICUs): intensive care units
(SD): standard deviation
(ANOVA): one-way analysis of variance
(ROC): receiver operating characteristic
(AUC): area under the curve
(SHAP): SHapley Additive exPlanations
(CITI): Collaborative Institutional Training Initiative

